# LEGAL AGE OF CONSENT FOR HIV TESTING AMONG ADOLESCENTS IN SUB SAHARAN AFRICA, A SYSTEMATIC REVIEW

**DOI:** 10.1101/2022.05.17.22275222

**Authors:** Getrud Joseph Mollel, Andrew Katende, Maryam Shahmanesh

## Abstract

Sub Saharan Africa (SSA) harbours more than 80% of adolescents living with HIV. High age of consent for HIV testing has been identified as one of the key barriers to adolescents’ access to HIV testing. We conducted a systematic literature review to demonstrate the status of age of consent policies in SSA and evidence of relationship between age of consent policies and adolescent’s uptake of HIV testing. We obtained peer reviewed literature from Medline, Embase, Scopus and Web of Science databases and policy review from national HIV testing guidelines and UNAIDS data reports. Age of consent for HIV testing in the region ranged between 12 and 18 years. Among 33 included countries, 14 (42.4%) had age of consent between 12 – 14 years, 9 (27.3%) had age of consent between 15 – 17 years and 10 countries (30.3%) still have the highest age of consent at 18 years as of 2019. Lowering age of consent has been associated with increased access to HIV testing among adolescents.

## INTRODUCTION

Sub Saharan Africa (SSA) harbours almost half of all HIV infected individuals and more than 80% of adolescents living with HIV.(1–3) AIDS related deaths have declined by more than a third globally, Eastern and Southern Africa showing the steepest reduction largely due to the scale up of HIV testing and coverage of antiretroviral therapy.(4) However, this improvement has not been uniform across all age groups. AIDS persisted as one of the leading causes of death for adolescents in Africa (5), with significantly higher rates of new infections in young girls than boys.(3) Reported reduction in AIDS related mortality among adolescents in 2018 was less than half the decline observed among adults aged above 20 years (16% versus 35%).(6)

Adolescence, defined as the age span from 10-19 years inclusively, is a complex developmental stage comprised of cognitive, physical, biological and psychological growth and characterised by social, personality and sexual exploration and maturation.(7,8) Adolescents have a higher likelihood of engaging in risky behaviours such as unsafe sexual practises which puts them at a higher risk of contracting HIV. Those who are part of HIV key populations including men who have sex with men, transgender people, intravenous drug users and commercial sex workers are even at a higher risk of contracting HIV and lower access to HIV testing especially in countries where they are criminalised.(5,9,10) Policies in many countries still restrict adolescents’ independent access to HIV testing without parental consent.(4) The proportion of adolescents who are aware of their HIV status is lower than that of adults.(11) Adolescent girls and young women contribute 26% of new HIV infections in Eastern and Southern Africa region, yet age of consent policies continue to be a barrier to HIV testing.(3,4) Furthermore, adolescents and youths represent the fastest growing population group in SSA which might precipitate the observed trend of new HIV infection among them, necessitating wider access to HIV testing and treatment services.(12)

Access to HIV testing is the gateway into HIV care and prevention. Therefore, as part of their commitment to scaling up of access to HIV services to all people including adolescents, WHO called upon heads of state to lower the age of consent for HIV testing.(13–15) Wider access to HIV testing among adolescents will complement several other interventions which are implemented in SSA with the aim of improving HIV outcomes among this age group. Such interventions include the adolescents’ friendly clinics, addressing HIV related stigma, promotion of abstinence and use of condoms, school based sex education, active case finding, use of incentives, provider initiated testing and counselling, voluntary male medical circumcision and other behavioural change interventions.(16–20) Adolescents require wider access to HIV testing especially for the realisation of public health benefits of starting antiretroviral treatment immediately after HIV diagnosis,(21) and other recent interventions such as Pre-exposure prophylaxis and HIV self-testing.(22,23) Removal of legal barriers to HIV testing could improve the extent to which adolescents benefit from these interventions.(24,25)

We conducted a systematic literature review on age of consent policies with an objective of demonstrating the status of age of consent policies in SSA and evidence of the relationship between age of consent and coverage of HIV testing among adolescents in the region.

## METHODS

We did a guideline review to summarise the age of consent for HIV testing in SSA followed by a systematic review of literature to assess the relationship between age of consent and access to HIV testing amongst adolescents in SSA. The PICO aspects of the review were as follows; population of interest was adolescents aged between 10-19 years, the intervention assessed was low age of consent, and the outcome was access to HIV testing among adolescents which was compared between countries with lower age of consent versus those with higher age of consent.

### Inclusion and exclusion criteria

We included the latest HIV testing guidelines supplemented with the UNAIDS global HIV reports and studies that were in the English language and from SSA countries. We defined SSA as countries geographically located South of the Sahara, as per the United Nations’ definition. We included studies that discussed the age of consent for HIV testing in adolescents in SSA region. Studies that included children, youths and young people were included in the initial search criteria due to the age overlap in definition of these age groups.

Adolescence is defined as the age between of 10 to 19 years inclusively, while a child is defined as any person from birth up to the age of 18 years, a young person is defined as the age of 10 up to 24 years, and a youth is any person between the ages of 15 to 24.(8,26). Studies that did not include adolescents were then excluded at later literature screening stages.

For the outcome of access to HIV testing we included studies that explored how age of consent influences HIV testing among adolescents aged between 10 to 19 years in SSA countries. We included studies which were conducted after 2013 as this was the year that WHO issued a review of consent age policies in SSA and called for governments consider lowering the age at which adolescents can independently access to HIV testing. (15,27) We did not limit studies based on their methodology. We included both quantitative and qualitative studies together with reviews of laws and policies related to age of consent for HIV testing. To assess changes in age of consent policies, we compared the age of consent reported in the 2013-WHO review to that stated in countries’ latest HIV testing guidelines and UNAIDS country data reports by 2019. We excluded studies if the focus was on age of consent for inclusion in HIV research.

### Search strategy

We searched for peer reviewed articles from Medline, Embase, Scopus and Web of science, and grey literature and policies from HIV testing guidelines, WHO guidelines and UNAIDS reports. *Table 1* summarises the key words, medical subject headings (Mesh) terms and proximity operators we used to search for the relevant literature.

**Table 1:** Search strategy.

|  |  |
| --- | --- |
| Search term 1 | "Informed consent" **OR** consent* **OR** "legal age" **OR** law* **OR** legislature **OR**<br>"Parental consent" |
| **AND** |  |
| Search term 2 | Adolescen* **OR** teen* **OR** youth* **OR** child* **OR** "young adult" **OR** "young<br>people" |
| **AND** |  |
| Search term 3 | (HIV adj2 test*) **OR** (HIV adj2 diagnos *) **OR** (HIV adj2 screen*) – *Medline and*<br>*embase proximity operator*<br>(HIV w/1 test*) **OR** (HIV w/1 diagnos*) **OR** (HIV w/1 screen*) - *Scopus proximity*<br>*operator*<br>(HIV NEAR/1 test*) **OR** (HIV NEAR/1 diagnos*) **OR** (HIV NEAR/1 screen*) –<br>*Web of science proximity operator* |
| **AND** |  |
| Search term 4 | "Africa South of the Sahara" **OR** "Sub-Saharan Africa*" **OR** "Subsaharan Africa*" **OR**<br>"Central Africa*" **OR** Cameroon* **OR** "Central African Republic*" **OR** Chad* **OR**<br>Congo* **OR** Gabon* **OR** "East* Africa*" **OR** Burundi* **OR** Djibouti* **OR** Eritrea* **OR**<br>Ethiopia* **OR** Kenya* **OR** Rwanda* **OR** Somalia* **OR** Sudan* **OR** Tanzania* **OR**<br>Uganda* **OR** "South* Africa*" **OR** Angola* **OR** Botswana* **OR** Lesotho* **OR** Malawi*<br>**OR** Mozambi* **OR** Namibia* **OR** Swazi* **OR** Eswatini **OR** Zambia* **OR** Zimbabwe* **OR**<br>"West* Africa*" **OR** "East* Africa*" **OR** Benin* **OR** "Burkina Faso*" **OR** "Cape<br>Verde*" **OR** "Cote d'Ivoire*" **OR** Gambia* **OR** Ghana* **OR** Guinea* **OR** Liberia* **OR**<br>Madagascar* **OR** Mali* **OR** Maurit* **OR** Niger* **OR** Senegal* **OR** "Sierra Leone*" **OR**<br>Togo* |

\* *Mesh terms*

### Literature screening

GJM conducted the literature search and screened all material according to the inclusion criteria. KA reviewed the materials against the inclusion and exclusion criteria. MS supervised literature search, screening and synthesis of results. Mendeley referencing software v.1.19.5 was used for literature management.

### Ethical considerations

We only included published materials without any inclusion of individual participants’ data. We therefore did not seek for further ethical approval

## RESULTS

We found 376, 446, 452 and 298 articles from Medline, Embase, Scopus and Web of science respectively. We obtained a total of 360 publications after removal of duplicates. We further excluded 261 and 76 articles at tittle screening and abstract screening stages respectively. 23 articles were eligible for full text screening where we further excluded 15 due to the following reasons; 2 articles discussed about consent for inclusion to HIV research and trials, full text literature for 1 article could not be found, 2 articles discussed HIV testing for children younger than 10 years old and 10 articles did not include discussion about age of consent among factors affecting HIV testing for adolescents. *Figure 1* shows the PRISMA flow diagram for literature screening. We finally included 8 articles from SSA. These included four quantitative studies,(28–31) one qualitative study,(32) and three reviews of HIV-specific policies, laws and legislative frameworks that affect adolescents’ access to HIV testing in SSA (33–35).

**Figure 1:**
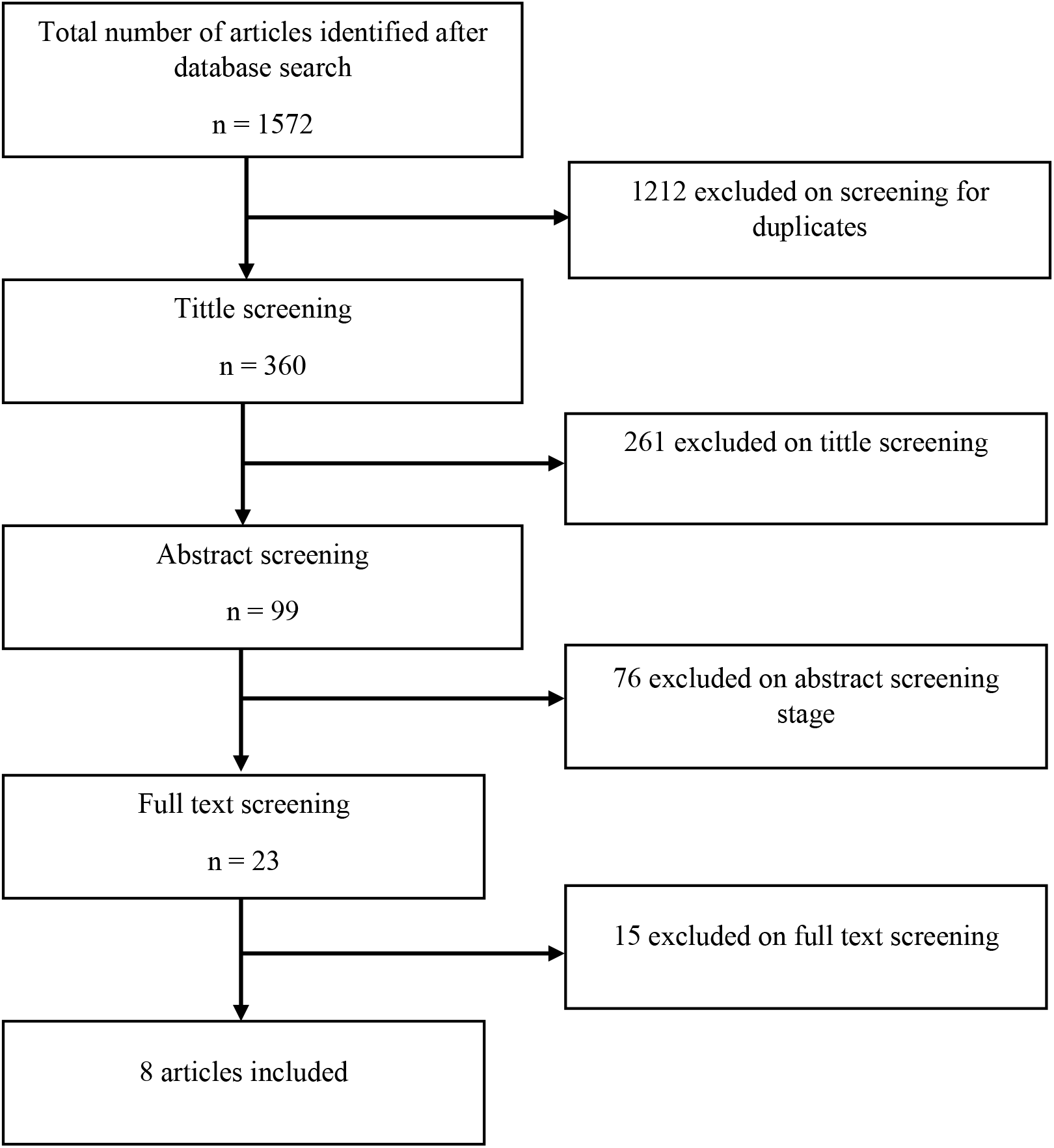
Prisma diagram.

We extracted age of consent from national HIV testing guidelines of Kenya, (36) Uganda,(37) Rwanda,(38) Ethiopia,(39) Malawi,(40) Zimbabwe,(41) Cameroon,(42) Ghana,(43) Swaziland,(44) Lesotho (45) and South Africa(46). We supplemented with a legal framework review from Tanzania (47) and UNAIDS country data reports of 2018 and 2019 (4,48). We included 33 SSA countries in total.

### Age of consent for HIV testing

The lowest age of consent for HIV testing among the countries included in this review was 12 years, while the highest was 18 years. *Table 2* shows the age of consent as reported in the WHO’s review of 2013 as compared to the country’s age of consent as indicated in the latest HIV testing guidelines or UNAIDS reports. In 2013, 5 (12.4%) countries had age of consent between 12 to 14 years, 15 to 17 years in 8 (24.2%) and 18 years in 12 (36.4%) countries. 6 (18.2%) countries had not defined age of consent for HIV testing for adolescents while 2 countries (Kenya and Botswana) were reported in 2013 as countries that apply maturity assessment by the healthcare provider rather than chronological age to determine adolescent’s ability to consent for HIV testing.(15). By 2019, the age of consent for HIV testing was 12 to 14 years in 14 (42.4%) countries, 15 to 17 years in 9 (27.3%) countries and 18 years in 10 (30.3%) countries. Between 2013 and 2019, Cameroon and Central African Republic lowered the age of consent from 18 years to 14 years, whereas Cote d’ivoire, Niger and Tanzania lowered it from 18 years to 16 years. This left Angola, Bukina Fasso, DRC, Djibouti, Ghana, Madagascar, Mali, Nigeria, Sierra Leone and Sudan with the highest age of consent for HIV testing in the region. The lowest age of consent in the region was 12 years in Uganda, South Africa, Rwanda, Lesotho and Swaziland. *Figure 2* shows a diagramatic presentation of changes in number of countries with respect to age of consent in 2013 as compared to 2019.

**Table 2:** Legal age of consent for HIV testing in SSA countries.

| Country | Age of consent<br>(years) reported in<br>2013<sup>(15)</sup> | Current age of consent (years) |
| --- | --- | --- |
| Angola | ND | 18 (4) |
| Benin | ND | 14 (4) |
| Botswana | NA | 16 (4) |
| Burkina Fasso | 18 | 18 (4) |
| Burundi | ND | 16 (48) |
| Cameroon | 18 | 14 (42) |
| Central African Republic | 18 | 14 (4) |
| Cote d ivoire | 18 | 16 (48) |
| DRC | 18 | 18 (48) |
| Djibouti | 18 | 18 (48) |
| Eswatini/Swaziland | 16 | 12 (44) |
| Ethiopia | 15 | 15 (39) |
| Ghana | 18 | 18 (43) |
| Kenya | NA | 15 (36) |
| Lesotho | 12 | 12 (45) |
| Liberia | 14 | 14 (4) |
| Madagascar | ND | 18 (4) |
| Malawi | 13 | 13 (40) |
| Mali | ND | 18 (4) |
| Mozambique | 16 | 14 (4) |
| Namibia | 16 | 14 (4) |
| Niger | 18 | 16 (4) |
| Nigeria | 18 | 18 (4) |
| Rwanda | 15 | 12 (38) |
| Senegal | 16 | 14 (4) |
| Sierra Leone | 18 | 18 (4) |
| South Africa | 12 | 12 (46) |
| Sudan | 18 | 18 (4) |
| Togo | ND | 14 (4) |
| Uganda | 12 | 12 (37) |
| United republic of Tanzania | 18 | 16 (47) |
| Zambia | 16 | 16 (4) |
| Zimbabwe | 16 | 16 (41) |

***ND- Age of minority with regard to HIV testing not defined***
***NA- Age is not applicable in criteria to decide adolescents who can consent for HIV testing, but maturity of the adolescents***

**Figure 2:**
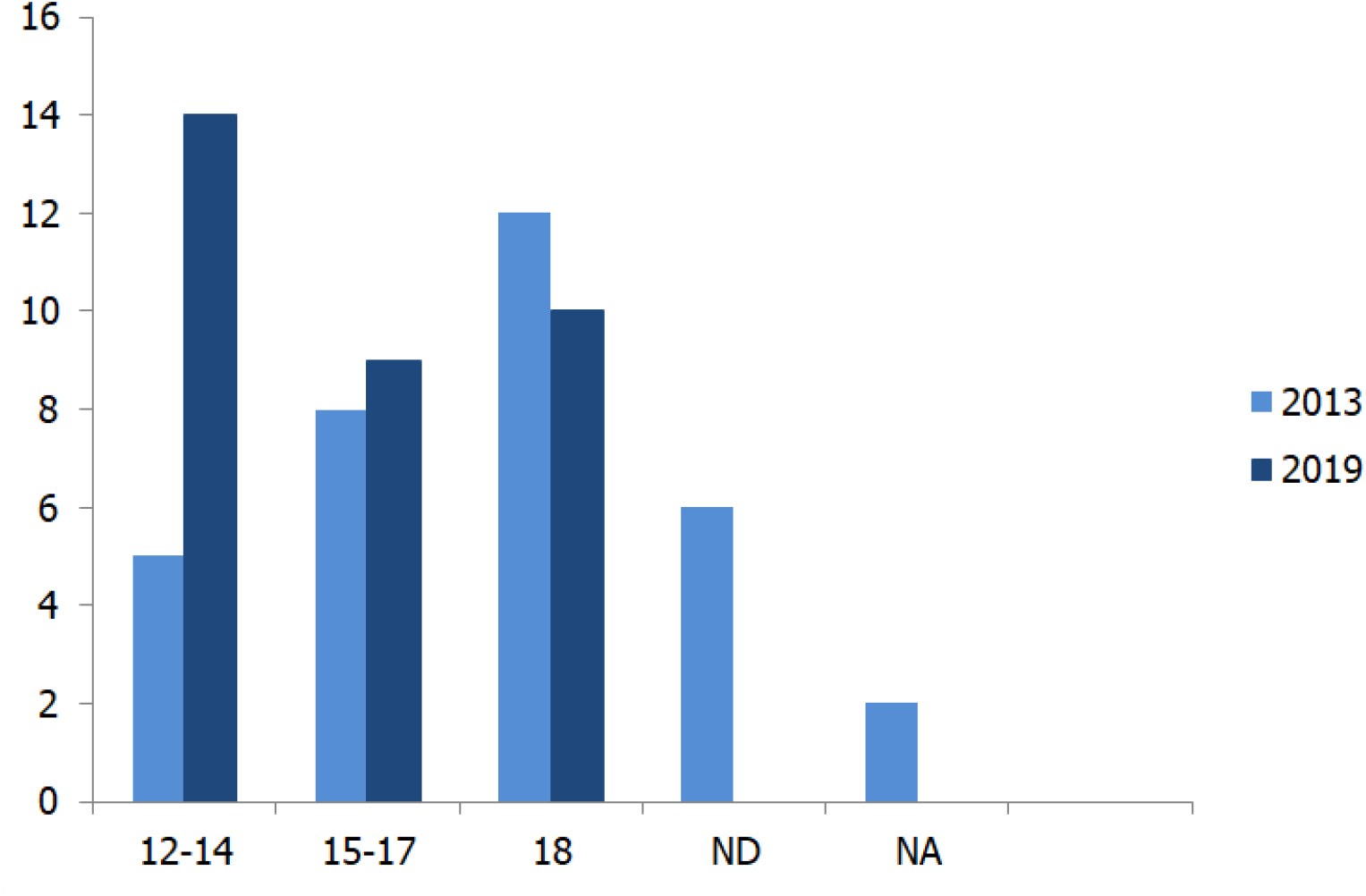
Age of consent in 2013 versus the age of consent in 2019. ND- Age of consent not defined, NA- Chronological age not applicable for consent

### Association between age of consent and HIV testing

McKinnon and colleagues quantitatively explored the association between age of consent and uptake of HIV testing among adolescents by using demographic and health survey data from 15 SSA countries. They found that age of consent below 16 years was associated with 11% (95%CI: 7.2-14.8) increase in uptake of HIV testing among adolescents.(28) Another study which used demographic and health survey data assessed predictors of HIV testing among adolescents and youths in Uganda, Congo, Mozambique and Nigeria also concluded that participants aged 20-24 years had higher odds of HIV testing than adolescents aged 15-19 years (aOR– 2.19; 99%CI 1.99-2.40). This finding was attributed to the fact that older participants could independently consent for HIV testing without requirement of parental consent. The overall uptake of HIV testing was low whereby only one third of the 23,367 included participants reported to have ever tested for HIV. They observed the highest coverage of HIV testing amongst adolescents in Uganda, where age of consent policies allow adolescents to access HIV testing from the age of 12 years.(29) These results are in congruence with adolescents’ views expressed in a qualitative assessment of HIV testing experience among adolescents, health care workers and parents in Kenya.(32) In this study, adolescents preferred to have independent access to HIV testing without parental consent requirements. They reported that they would prefer post-test counselling support and a choice on how to handle disclosure of their HIV status. While adolescents’ opinion should be taken into consideration when designing policies to improve their uptake of HIV testing services, parents on the other hand preferred to be informed about HIV status of their children even when it’s contrary to adolescents’ preference.(32) Mixed findings were also reported from cross sectional studies aimed at assessing parents’ attitudes towards implementation of HIV testing at schools in South Africa following findings of a high desire for a school-based HIV testing services among adolescents.(30,31) Among 801 parents who participated in the survey, 46% reported that students can access HIV testing at school without requirements of parental consent, whereas 39% reported that HIV results should be communicated to the adolescents in the presence of their parents.(30) Healthcare workers in Kenya also reported that they sometimes deny adolescents testing services unless they are pregnant, married or parents, (32) despite the fact that the HIV testing guideline in Kenya allow adolescents aged above 15 years to access HIV testing without parental consent.(36)

### Policies and legal frameworks in relation to HIV testing

We included three articles that discussed policies, laws and legal frameworks affecting adolescents’ access to HIV testing in SSA. These included one desk review of HIV related laws of Angola, Benin, Burkina Fasso, Burundi, Cape Verde, Central African Republic, Chad, Comoros, Congo, Cote d’Ivoire, Democratic Republic of Congo, Equatorial Guinea, Gambia, Guinea, Guinea Bissau, Kenya, Liberia, Madagascar, Mali, Mauritania, Mauritius, Mozambique, Niger, Senegal, Sierra Leone, Tanzania, Togo and Uganda,(33) a review of health policy gaps that affect adolescents access to HIV services in Rwanda,(34) and a review of South African legislative reform for improvement of adolescents’ access to sexual and reproductive health services including HIV testing.(35) The review from 28 SSA countries examined HIV related laws against WHO’s recommendations to lower the age of consent for HIV testing and ensure access to HIV testing, counselling and treatment among adolescents. Of the reviewed laws, 11 countries had explicitly stated the age at which adolescents can independently access HIV testing whereby only 7 of them had the age of consent set below 18 years. Criteria for independent access to HIV testing for HIV testing other than chronological age included sufficient maturity, emancipated minor, pregnant or married adolescents, adolescents who are married or those at higher risk of acquiring HIV infection for Kenya, Comoros, Mauritius, Madagascar, Togo, Sierra Leone and Niger. None of the laws had an explicit definition of maturity with respect to HIV testing. Age of consent for HIV testing in Kenya HIV testing policies was found to be different from the age stated in the law. This review highlighted the need of reforms to address inadequacy of HIV specific laws in SSA in enhancing adolescents’ independent access to HIV testing.(33) Additionally, Binagwaho and his colleagues used a human right approach to argue that adolescents have a right to access confidential HIV testing services and their results to only be disclosed if they agree to it. They called for a reduction of age of consent for HIV testing from 21 years which was a default age at which an adolescent acquired majority and ability to consent for HIV testing in Rwanda to coincide with adolescents sexual behaviours.(34) Current HIV testing guidelines in Rwanda allows for independent consent for HIV testing for adolescents from the age of 12 years and mature enough to understand implications of HIV testing and to be able to cope in case of positive results.(38) Through its legislative reforms, South Africa also allowed independent consent for HIV testing from mature adolescents aged 12 years and above.(35) However, adolescents who are above 12 years and rendered by healthcare provider to be not mature enough to understand the benefits of HIV testing are still required by the South African HIV testing guideline to provide parental consent.(46)

## DISCUSSION

We found that the majority of the countries have the age of consent for HIV testing between 12 and 14 years and all countries have explicitly mentioned the age at which adolescents can independently access HIV test. This is substantial progress compared to 2013 where 6 countries had not defined the age of consent for HIV testing. Access to HIV testing is an entry point to HIV treatment services and prevention of further new infection and the limited data in this review suggests that age of consent is associated with uptake of HIV testing amongst adolescents. However, despite progress in removing age of consent as a barrier towards HIV testing, adolsecents in the region still contribute more than one third of new HIV infections.(3)

There were only two studies that quantitatively assessed the association of HIV testing and legal age of consent where a higher uptake was reported among countries with low age of consent (28,29). Suboptimal response of countries may be attributed to paucity of evidence showing the extent to which adolescents’’ HIV testing improves after lowering age of consent. A study among adolescents from 32 states of the US where requirement of parental consent for adolescents to access HIV testing was waived reported that there was no significant statistical association between legal ability to consent and uptake of HIV testing (aOR-0.3% (95%CI 0.1,1.1)).(49) However the results could have been swayed by several limitations of the study including recall and social desirability bias and default exclusion of non-internet users who could have been systematically different from internet using adolescent who participated in the study. Arguably, everyone including adolescents have rights to access HIV testing services especially now when universal access to health care is among the priority global health agendas in developing countries and globally. However, it is necessary to understand the added advantage of lowering the age of consent together with establishing the role of contextual factors that might limit adolescents’ access to HIV testing even in the settings of low age of consent.

Secondly, there are no evidence based maturity assessment methods to determine adolescents who should be allowed to access HIV testing without parental consent. Most national guidelines point towards the use of sexual maturity such as being pregnant, being a parent or in a higher risk of contracting HIV as a gateway for independent access to HIV testing among adolescents. The use of sexual related indicators of maturity systematically excludes adolescents who are not sexually active but could have been perinatally infected-long term slow progressors.(50,51) It also segregates against adolescents who are not yet sexually active but may benefit from HIV testing as a gateway into HIV prevention counseling. Furthermore, such criteria provide independent access to HIV testing after exposure to the risk of infection.

Additionally, we found that it is largely left at health workers’ individual discretion to decide whether adolescents are mature enough to access HIV testing. Definition of maturity is bound to differ between individuals, and across cadres i.e., nurses, medical doctors, social scientists, psychologists, lawyers and counsellors. It may also differ depending on social construct, culture and development of both the provider and the client.(52) Adolescent health experts from various fields including ethics, sociology, law and medicine who convened in 2015 recommended assessment of adolescent’s capability for decision making in clinical care should include understanding of the legal requirements, conducive and non-judgemental environment and objective assessment of social, emotional and cognitive development.(53) Guidelines for evaluation of adolescents capacity to consent for HIV testing could also be adopted from “Gillick competence” assessment criteria which originated from the United Kingdom, used mainly to assess adolescents cognitive ability to consent for a variety medical services such as sexual health service and immunization.(54,55) Countries in SSA could adopt and validate such kind of assessments which would enable timely HIV diagnosis among adolescents. It would also give access to adolescents who may not have been exposed already, and open the window for HIV prevention counselling to them and their peers.

Studies showed that parents required to be notified about adolescents’ HIV status despite of their disclosure preferences which contributes to healthcare workers hesitancy in offering HIV test for adolescents even in settings of favourable consent policies.(30,32) Unwanted and uninformed disclosure to third parties including parents might discourage adolescents from accessing HIV testing service even after lowering the legal age of consent. Parents’ and healthcare workers should therefore be sensitized on importance of lowering the age of consent to allow truly autonomous access of HIV testing among adolescents. Furthermore, social networks including families, peers and surrounding societies may play an important demand-side influence on adolescent’s decision and ability to seek for HIV testing.(56) Their perceptions on adolescents’ autonomous access to HIV testing should therefore be explored and addressed in order to maximise benefits of lower age of consent on adolescents’ access to HIV testing.

This review adds to the existing literature that calls for lowering the age of consent for HIV testing in SSA region while assessing and addressing other factors that might impede benefits of lower age of consent. It has triangulated findings from legal reviews, HIV testing guidelines and qualitative and quantitative studies. This review was not without limitations. We did not include articles and national guidelines which were not published in English which may bias the results. We also included only 33 countries from the region. However, this was done based on the countries which were included in the WHO 2013 report to allow comparison, and availability of data on age of consent policies.

## CONCLUSION

Age of consent for HIV in SSA region ranges from 12 to 18 years. Majority of countries in SSA countries currently allow adolescents aged between 12 to 14 years to independently consent for HIV testing. However, there were countries that continued to maintain the highest age of consent (18 years). There is a need for context specific evaluation of benefits of lower age of consent and other factors that impede adolescents’ access to HIV testing even in settings of favourable consent policies.

## Data Availability

There is no primary data collected for completion of this work

## Contributions from authors

GJM conceived the idea, formulated the research question, conducted literature search, screening of materials and drafted the manuscript

KA reviewed the literature search against eligibility criteria and reviewed the manuscript. MS supervised the literature search and reviewed the manuscript

